# Longitudinal assessment of diagnostic test performance over the course of acute SARS-CoV-2 infection

**DOI:** 10.1101/2021.03.19.21253964

**Authors:** Rebecca L. Smith, Laura L. Gibson, Pamela P. Martinez, Ruian Ke, Agha Mirza, Madison Conte, Nicholas Gallagher, Abigail Conte, Leyi Wang, Rick Fredrickson, Darci C. Edmonson, Melinda E. Baughman, Karen K. Chiu, Hannah Choi, Tor W. Jensen, Kevin R. Scardina, Shannon Bradley, Stacy L. Gloss, Crystal Reinhart, Jagadeesh Yedetore, Alyssa N. Owens, John Broach, Bruce Barton, Peter Lazar, Darcy Henness, Todd Young, Alastair Dunnett, Matthew L. Robinson, Heba H. Mostafa, Andrew Pekosz, Yukari C. Manabe, William J. Heetderks, David D. McManus, Christopher B. Brooke

## Abstract

**What is already known about this topic?:** Diagnostic tests and sample types for SARS-CoV-2 vary in sensitivity across the infection period.

**What is added by this report?:** We show that both RTqPCR (from nasal swab and saliva) and the Quidel SARS Sofia FIA rapid antigen tests peak in sensitivity during the period in which live virus can be detected in nasal swabs, but that the sensitivity of RTqPCR tests rises more rapidly in the pre-infectious period. We also use empirical data to estimate the sensitivities of RTqPCR and antigen tests as a function of testing frequency.

**What are the implications for public health practice?:** RTqPCR tests will be more effective than rapid antigen tests at identifying infected individuals prior to or early during the infectious period and thus for minimizing forward transmission (provided results reporting is timely). All modalities, including rapid antigen tests, showed >94% sensitivity to detect infection if used at least twice per week. Regular surveillance/screening using rapid antigen tests 2-3 times per week can be an effective strategy to achieve high sensitivity (>95%) for identifying infected individuals.

## INTRODUCTION

Frequent rapid diagnostic testing is critical for restricting community spread of SARS-CoV-2 by allowing the timely identification and isolation of infected individuals to interrupt the chain of transmission. Quantitative reverse transcription polymerase chain reaction (RTqPCR)-based detection of viral RNA within nasal swab or saliva samples represents the gold standard for sensitivity in detecting the presence of SARS-CoV-2, yet supply shortages, cost, and infrastructure limitations have made it difficult to achieve high testing frequency and volume with the rapid reporting of results needed to mitigate transmission effectively.

Recently, there has been considerable interest in the potential of rapid antigen tests to expand diagnostic testing capacity due to the ease of use, availability, cost, and rapid time-to-results^1^. However, data for their use in screening asymptomatic individuals is sparse. Enthusiasm for their widespread deployment has been further tempered by well-publicized examples of false positive results in people with low pre-test probability of infection, and by reports suggesting they lack sensitivity compared with RTqPCR, potentially making them less effective at mitigating community spread^2,3^.

Here, we compare the sensitivities of nasal and saliva RTqPCR tests with the Quidel Sofia SARS Antigen Fluorescent Immunoassay (FIA) over the course of mild or asymptomatic acute SARS-CoV-2 infection through daily sampling of individuals enrolled early during infection.

## METHODS

This study was approved by the Western Institutional Review Board, and all participants consented freely.

### Participants

All on-campus students and employees of the University of Illinois at Urbana-Champaign are required to submit saliva for RTqPCR testing every 2-4 days as part of the SHIELD campus surveillance testing program. Those testing positive are instructed to isolate, and were eligible to enroll in this study for a period of 24 hours following receipt of their positive test result. Close contacts of individuals who test positive (particularly those co-housed with them) are instructed to quarantine and were eligible to enroll for up to 5 days after their last known exposure to an infected individual. All participants were also required to have received a negative saliva RTqPCR result 7 days prior to enrollment.

Individuals were recruited via either a link shared in an automated text message providing isolation information sent within 30 minutes of a positive test result, a call from a study recruiter, or a link shared by an enrolled study participant or included in information provided to all quarantining close contacts. In addition, signs were used at each testing location and a website was available to inform the community about the study.

Participants were required to be at least 18 years of age, have a valid university ID, speak English, have internet access, and live within 8 miles of the university campus. After enrollment and consent, participants completed an initial survey to collect information on demographics and health history, including suspected date of SARS-CoV-2 exposure. They were then provided with sample collection supplies.

Participants who tested positive prior to enrollment or during quarantine were followed for up to 14 days. Quarantining participants who continued to test negative by saliva RTqPCR were followed for up to 7 days after their last exposure. All participants’ data and survey responses were collected in the Eureka digital study platform.

### Sample collection

Each day, participants were remotely observed by study staff collecting:

1. 2 mL of saliva into a 50mL conical tube.
2. 1 nasal swab from a single nostril using a foam-tipped swab that was placed within a dry collection tube.
3. 1 nasal swab from the other nostril using a flocked swab that was subsequently placed in a collection vial containing viral transport media (VTM).

The order of nostrils (left vs. right) used for the two different swabs was randomized. For nasal swabs, participants were instructed to insert the soft tip of the swab at least 1 cm into the indicated nostril until they encountered mild resistance, rotate the swab around the nostril 5 times, leaving it in place for 10-15 seconds. After daily sample collection, participants completed a symptom survey. A courier collected all participant samples within 1 hour of collection using a no-contact pickup protocol designed to minimize courier exposure to infected participants.

### Saliva RTqPCR

After collection, saliva samples were stored at room temperature and RTqPCR was run within 12 hours of initial collection. The protocol for direct saliva-to-RTqPCR assay used has been detailed previously^4^. In brief, saliva samples were heated at 95°C for 30 minutes, followed by the addition of 2X TBS at a 1:1 ratio (final concentration 1X TBE) and Tween-20 to a final concentration of 0.5%. Samples were assayed using the Thermo Taqpath COVID-19 assay.

### Quidel assay

Foam-tipped nasal swabs were placed in collection tubes and stored at 4°C overnight based on guidance from the manufacturer. The morning after collection, swabs were run through the Sofia SARS antigen FIA on Sofia 2 devices according to the manufacturer’s protocol.

### Nasal swab RTqPCR

Collection tubes containing VTM and flocked nasal swabs were stored at −80°C after collection and were subsequently shipped to Johns Hopkins University for RTqPCR and virus culture testing. After thawing, VTM was aliquoted for RTqPCR and infectivity assays. One ml of VTM from the nasal swab was assayed on the Abbott Alinity per manufacturer’s instructions in a College of American Pathologist and CLIA-certified laboratory.

### Nasal virus culture

VeroTMPRSS2 cells were grown in complete medium (CM) consisting of DMEM with 10% fetal bovine serum (Gibco), 1 mM glutamine (Invitrogen), 1 mM sodium pyruvate (Invitrogen), 100 U/ml of penicillin (Invitrogen), and 100 μg/ml of streptomycin (Invitrogen)^5^. Viral infectivity was assessed on VeroTMPRSS2 cells as previously described using infection media (IM; identical to CM except the FBS is reduced to 2.5%)^6^. When a cytopathic effect was visible in >50% of cells in a given well, the supernatant was harvested. The presence of SARS-CoV-2 was confirmed through RTqPCR as described previously by extracting RNA from the cell culture supernatant using the Qiagen viral RNA isolation kit and performing RTqPCR using the N1 and N2 SARS-CoV-2-specific primers and probes in addition to primers and probes for human RNaseP gene using synthetic RNA target sequences to establish a standard curve^7^.

### Data Analysis

At the time of analysis, nasal samples from 30 participants had been analyzed by virus culture and RTqPCR. Therefore, analyses that consider either nasal RTqPCR or viral culture results were conducted based on a limited participant set. All confidence intervals around sensitivity were calculated using binconf from the Hmisc package in R version 3.6.2.

The sensitivity of each of the tests was analyzed in three different ways:

First, the ability of each test (antigen, saliva RTqPCR, or nasal RTqPCR) to detect an infected person on a particular day relative to the day of first positive viral culture (“daily sensitivity”) was calculated. Daily sensitivity was not calculated for timepoints with fewer than 5 observed person-days.

Second, the ability of each test to detect an infected person according to their viral culture status (“status sensitivity”) was calculated. Viral culture status was defined as “pre-positive” on days prior to the first positive viral culture result, “positive” on days for which viral culture results were positive, and “post-positive” on days with negative viral culture results that occur after the first positive culture result. Status sensitivity was defined as the proportion of person-days with a positive result.

Finally, we calculated the ability of repeated testing over a 14-day period to detect an infected person (“protocol sensitivity”) using a value-of-information approach. Seven different testing frequencies were considered: daily, every other day, every third day, and so on, up to weekly sampling. For each individual, the result of testing on a given schedule was calculated for each potential starting date, with test results interpreted in parallel (all tests must be negative to be considered negative). For instance, each person contributed two observations to the “every other day” schedule, one starting on the first day of the study and the other starting on the second day of the study. The proportion of “observations” with a positive result (at least one positive test in the sampling timeframe) was considered to be the sensitivity of that testing protocol (test and frequency combination).

All code used in analyses can be found here: https://github.com/rlsdvm/CovidDetectAnalysis

## Results

Table 1 shows demographic information for study participants reported here. The majority of participants (21/30, 70%) were non-Hispanic white and the average age was 32.50 (SD 12.29).

**Table 1:** Demographic information on participants enrolled in the COVID detect study.

| Variable |  | Data |
| --- | --- | --- |
|  |  | n=30 |
| Weight (mean (SD)) |  | 176.00 (51.17) |
| Height in inches (mean (SD)) |  | 67.57 (4.94) |
| Age (mean (SD)) |  | 32.50 (12.29) |
| Race (%) | Native American | 0 ( 0.0) |
|  | Asian | 1 ( 3.3) |
|  | Black | 2 ( 6.7) |
|  | Other | 3 (10.0) |
|  | Pacific Islander | 0 ( 0.0) |
|  | White | 24 (80.0) |
| Gender (%) | Female | 12 (40.0) |
|  | Male | 18 (60.0) |
| Ethnicity (%) | Hispanic | 6 (20.0) |
|  | Non-Hispanic | 24 (80.0) |

We first estimated the daily sensitivities of nasal and saliva RTqPCR and antigen tests relative to the day of first nasal swab viral culture positivity, which was used as a surrogate marker of infectious virus shedding (**Table 2, Figure 1**). We also used the viral culture data to measure the status sensitivities of each test before, during, and after viral shedding (**Figure 2**).

**Table 2:** Daily sensitivity of each test platform by day relative to the day of first nasal swab viral culture positivity.

| Days before<br>(-1,-2), on (0), or<br>after the day of<br>first positive<br>culture | Antigen |  | Saliva RTqPCR |  | Nasal RTqPCR |  | Total |
| --- | --- | --- | --- | --- | --- | --- | --- |
|  | Daily<br>Sensitivity | Number<br>positive | Daily<br>Sensitivity | Number<br>positive | Daily<br>Sensitivity | Number<br>positive |  |
| -2 | 0.333 | 2 | 0.833 | 5 | 0.667 | 4 | 6 |
| -1 | 0.455 | 5 | 0.636 | 7 | 0.727 | 8 | 11 |
| 0 | 0.875 | 21 | 0.958 | 23 | 1.000 | 24 | 24 |
| 1 | 0.960 | 24 | 1.000 | 25 | 1.000 | 25 | 25 |
| 2 | 0.960 | 24 | 0.960 | 24 | 1.000 | 25 | 25 |
| 3 | 0.920 | 23 | 0.920 | 23 | 1.000 | 25 | 25 |
| 4 | 0.760 | 19 | 0.960 | 24 | 1.000 | 25 | 25 |
| 5 | 0.640 | 16 | 0.840 | 21 | 0.960 | 24 | 25 |
| 6 | 0.560 | 14 | 0.920 | 23 | 0.880 | 22 | 25 |
| 7 | 0.250 | 6 | 0.667 | 16 | 0.792 | 19 | 24 |
| 8 | 0.182 | 4 | 0.682 | 15 | 0.909 | 20 | 22 |
| 9 | 0.045 | 1 | 0.500 | 11 | 0.727 | 16 | 22 |
| 10 | 0 | 0 | 0.500 | 10 | 0.900 | 18 | 20 |
| 11 | 0.05 | 1 | 0.500 | 10 | 0.800 | 16 | 20 |
| 12 | 0 | 0 | 0.368 | 7 | 0.526 | 10 | 19 |
| 13 | 0 | 0 | 0.231 | 3 | 0.385 | 5 | 13 |

**Figure 1:**
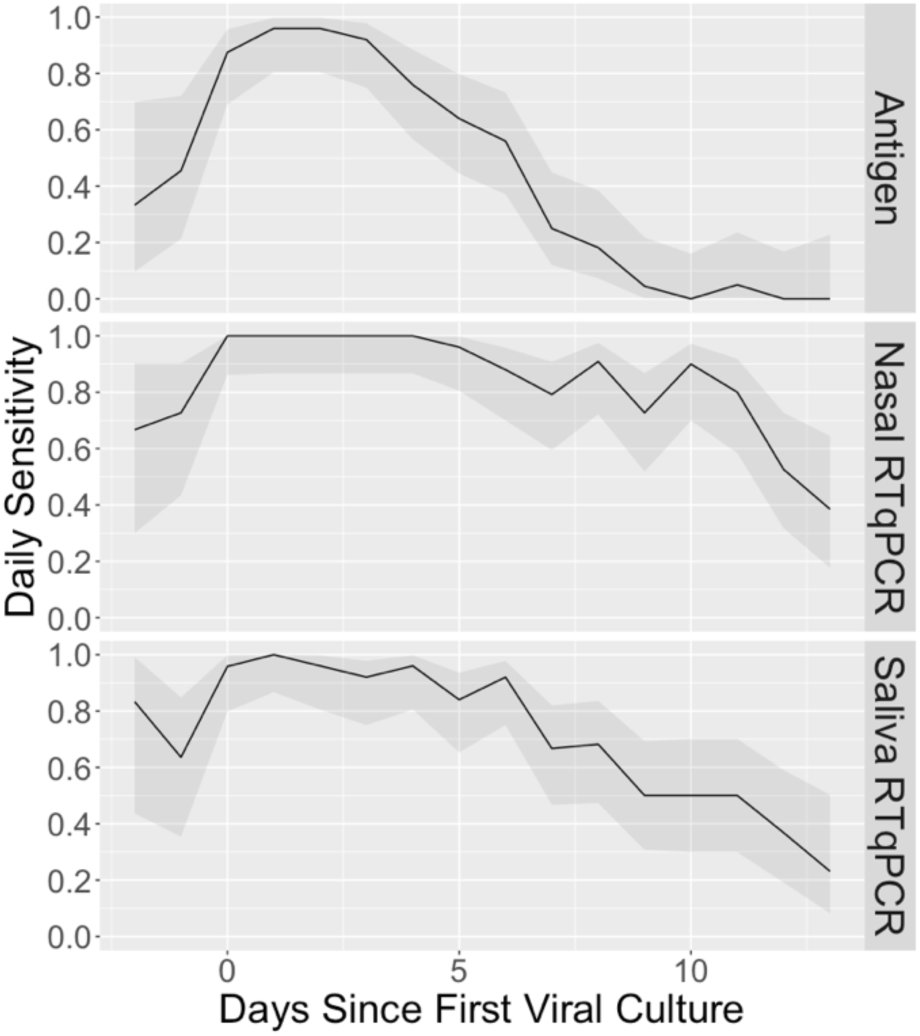
Daily sensitivity of each test platform by day relative to the day of first positive viral culture result. Shaded areas represent the 95% confidence interval around the observed proportion.

**Figure 2:**
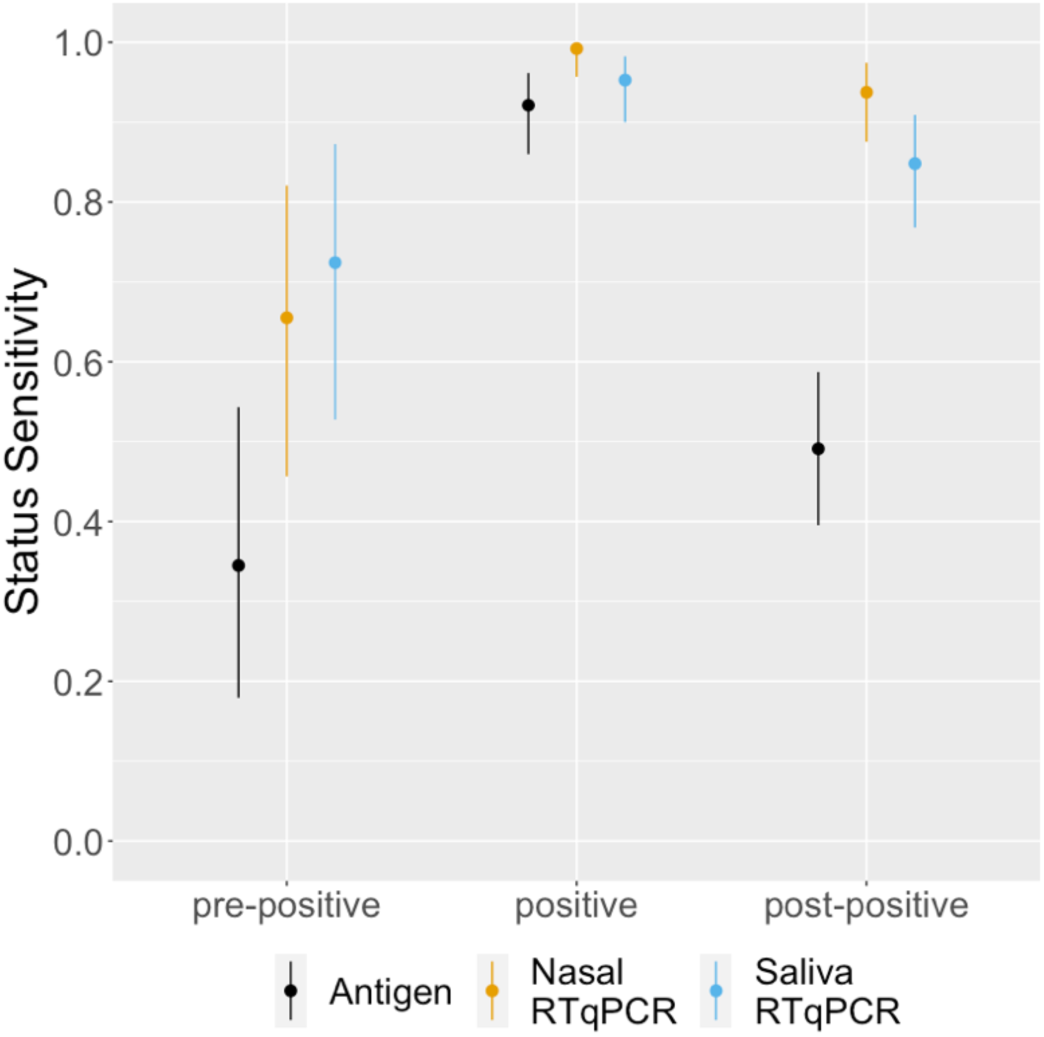
Status sensitivity of each test platform relative to viral culture positivity. Bars indicate the 95% confidence interval around the observed proportion. Pre-positive (n=29) refers to samples taken on days before the first viral culture-positive sample collected from each individual. Positive (n=127) refers to samples taken on days for which viral culture results were positive. Post-positive (n=112) refers to samples taken on days with negative viral culture results that occur after the first positive culture result.

Prior to the first day of detectable shedding of infectious virus, both RTqPCR tests had higher daily sensitivity (0.706 for both saliva and nasal) than the antigen test (0.412). For all three tests, daily and status sensitivity peaked during days in which infectious virus shedding was detectable, as would be expected. Antigen test daily sensitivity declined precipitously after infectious virus could no longer be detected in nasal swabs, dropping below 0.5 within a week after the onset of culture positivity, while both nasal and saliva RTqPCR platforms only showed minor decreases in sensitivity, remaining at 0.792 and 0.667 after a week, respectively.

We next estimated the protocol sensitivities, or how the ability of each of test platform to detect infected individuals was affected by differences in testing frequencies (**Table 3, Figure 3**). Protocol sensitivity was defined at the schedule level, where the numerator is the number of testing schedules resulting in at least one positive test and the denominator is the number of testing schedules examined, where a testing schedule is defined as a set of samples from one participant taken at a given frequency. In **Figure 3**, we calculated the effects of varying testing frequency on sensitivity to detect infected individuals on days where nasal swabs were viral culture positive in the top panel. In the bottom panel of **Figure 3**, we examined sensitivity to detect infected individuals at any stage of infection.

**Table 3:**
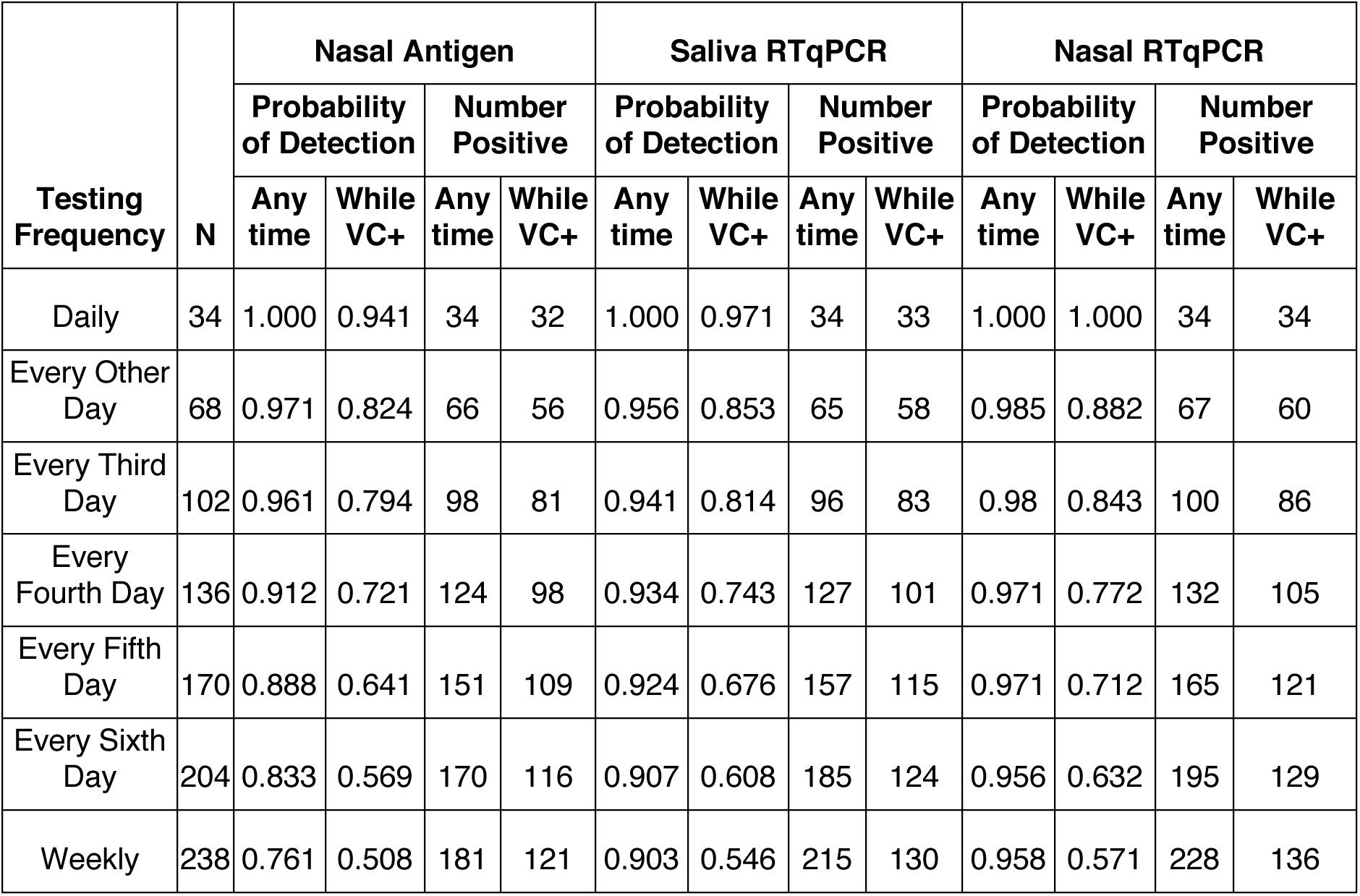
Protocol sensitivity of each test platform to detect an infected person during a 14-day testing period, relative to the frequency of testing. “Any time” refers to detection of the individual at any point in the 14-day testing period; “While VC+” refers to detection of the individual before or during the time in which their viral culture was positive.

**Figure 3:**
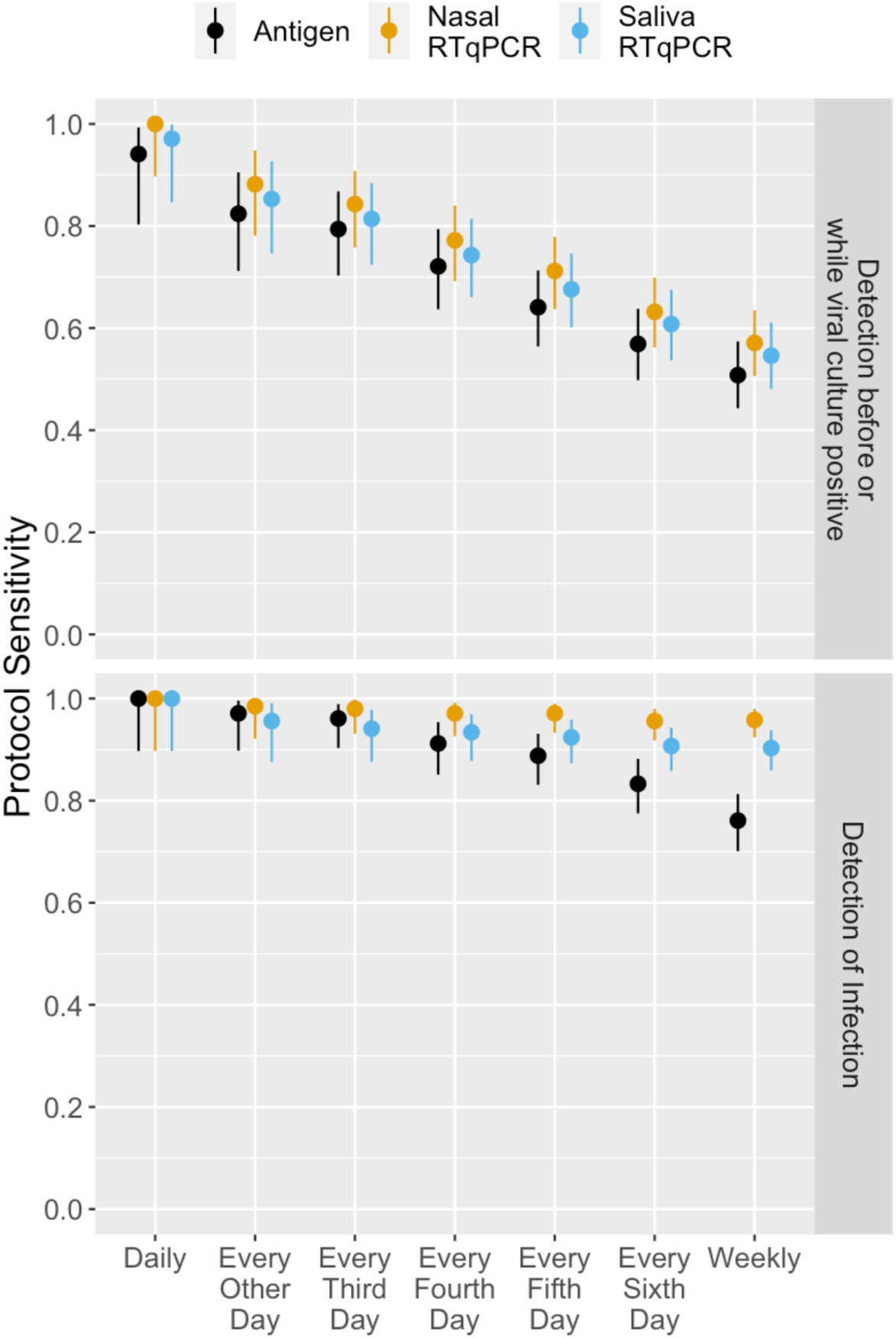
Protocol sensitivity of each test platform to detect an infected person **(top)** before or during days where nasal samples were viral culture positive or **(bottom)** at any time, over a 14-day testing period, relative to frequency of testing. Lines indicate 95% confidence interval around the observed proportion.

## Discussion

Our data demonstrate that the sensitivities of RTqPCR and antigen tests vary significantly over the course of SARS-CoV-2 infection. Prior to the presumed infectious period (here defined as the period during which infectious virus could be detected in nasal swab samples), the daily sensitivities of nasal and saliva RTqPCR tests were higher than that of the Quidel Sofia SARS Antigen FIA, suggesting that RTqPCR tests will be more effective at identifying infected individuals before they transmit to others.

Both RTqPCR and antigen tests peak in daily and status sensitivities when infectious virus is detectable in nasal swab samples, suggesting that all three modalities can be effective at identifying individuals during the presumed infectious period. After this period, the daily sensitivity of RTqPCR tests decreased gradually, with saliva RTqPCR dropping faster than nasal RTqPCR. These dynamics are consistent with those described previously for RTqPCR^8,9^. In contrast, the daily sensitivity of the antigen test declined very quickly, suggesting that this test will be less effective at identifying individuals during later stages of infection. This may limit diagnosis and contact-tracing efforts in test-limited environments.

Previous studies have suggested that frequent testing would maximize the ability of a given test modality to detect infected individuals^10,11^. We found that all testing modalities showed almost 95% protocol sensitivity to detect infection if used at least twice per week. When applied weekly, protocol sensitivity remained very high for nasal RTqPCR, declined slightly to 90% for saliva RTqPCR, and dropped to only 76% for the antigen test.

When we compared the abilities of different testing frequencies to identify individuals while infectious virus was detectable in nasal samples, we observed a clear reduction in protocol sensitivity for all testing modalities when testing frequencies decreased below daily. The reduction in protocol sensitivity was most pronounced for the antigen test, which dropped to 0.72 with testing every fourth day, however, both RTqPCR tests were only slightly better at 0.74 (saliva) and 0.77 (nasal). Altogether, these data demonstrate the importance of frequent testing regardless of test modality for identifying individuals while they are contagious.

This is the first study to compare the longitudinal performance of rapid antigen and RTqPCR tests with infectious virus shedding in a well-defined population early in SARS-CoV-2 infection. We found that all three diagnostic tests demonstrated a high degree of daily sensitivity during the presumed infectious period, but that the RTqPCR tests exhibited superior daily sensitivities prior to this period. Our data suggest that RTqPCR tests can be more effective than antigen tests at mitigating community spread of SARS-CoV-2, but only if the turnaround time for RTqPCR results is short. Finally, these data also quantitatively demonstrate the importance of frequent (at least twice per week) screening to maximize likelihood of detecting infected individuals regardless of testing modality.

## Data Availability

All code used in analyses can be found here: https://github.com/rlsdvm/CovidDetectAnalysis

## Acknowledgments

This study was funded by the NIH RADx-Tech program under 3U54HL143541-02S2. The views expressed in this manuscript are those of the authors and do not necessarily represent the views of the National Institute of Biomedical Imaging and Bioengineering; the National Heart, Lung, and Blood Institute; the National Institutes of Health, or the U.S. Department of Health and Human Services. Sofia 2 devices and associated supplies were provided to Carle Foundation Hospital by Quidel, however Quidel played no role in the design of the study or the interpretation or presentation of the data.

We also thank Shumon Ahmed, Carly Bell, Nate Bouton, Callie Brennen, Justin Brown, Coleco Buie, Emmaline Cler, Gary Cole, Trey Coleman, Lauren Engels, Savannah Feher, Kelsey Fox, Lexi Freeman, Yesenia Gonzalez, Montez Harris, Dan Hiser, Ayeshah Hussain, Daryl Jackson, Michael Jenkins, Kalombo Kalonji, Syntyche Kanku, Steven Krauklis, Mary Krouse, Elmore Leshoure, Joe Lewis, Angel Lopez, Guadalupe Lopez, Emily Luna, Chun Huai Luo, Colby Mackey, Skyler McLain, Yared Berhanu Melesse, Madison O’Donnell, Savanna Pflugmacher, Denver Piatt, Skyler Pierce, Jessica Quicksall, Gina Quitanilla, Ameera Samad, MacKenzie Scroggins, Monique Settles, Macie Sinn, Pete Varney, Evette Vlach, and Raeshun Williams-Chatman for their efforts supporting recruitment, enrollment, logistics, and sample collection. We also thank Jeffrey Olgin, Noah Peyser, and Xochitl Butler for assistance with the Eureka platform, Michelle Lore for assistance with REDcap, and Gillian Snyder for assistance in development of study protocols and logistics.

